# High frequency of artemisinin partial resistance mutations in the great lake region revealed through rapid pooled deep sequencing

**DOI:** 10.1101/2024.04.29.24306442

**Authors:** Neeva Wernsman Young, Pierre Gashema, David Giesbrecht, Tharcisse Munyaneza, Felicien Maisha, Fred Mwebembezi, Rule Budodo, Alec Leonetti, Rebecca Crudale, Vincent Iradukunda, Ntwari Jean Bosco, Ross M. Boyce, Celine I. Mandara, Grace K. Kanyankole, Edgar Mulogo, Deus S. Ishengoma, Stan Hangi, Corine Karema, Jean-Baptiste Mazarati, Jonathan J Juliano, Jeffrey A. Bailey

## Abstract

In Africa, the first *Plasmodium falciparum* Kelch13 (K13) artemisinin partial resistance mutation 561H was first detected and validated in Rwanda. Surveillance to better define the extent of the emergence in Rwanda and neighboring countries as other mutations arise in East Africa is critical. We employ a novel scheme of liquid blood drop preservation combined with pooled sequencing to provide a cost-effective rapid assessment of resistance mutation frequencies at multiple collection sites across Rwanda and neighboring countries. Malaria-positive samples (n=5,465) were collected from 39 health facilities in Rwanda, Uganda, Tanzania, and the Democratic Republic of the Congo (DRC) between May 2022 and March 2023 and sequenced in 199 pools. In Rwanda, K13 561H and 675V were detected in 90% and 65% of sites with an average frequency of 19.0% (0-54.5%) and 5.0% (0-35.5%), respectively. In Tanzania, 561H had high frequency in multiple sites while it was absent from the DRC although 675V was seen at low frequency. Conceringly candidate mutations were observed: 441L, 449A, and 469F co-occurred with validated mutations suggesting they are arising under the same pressures. Other resistance markers associated with artemether-lumefantrine are common: *P. falciparum* multidrug resistance protein 1 N86 at 98.0% and 184F at 47.0% (0-94.3%) and *P. falciparum* chloroquine resistance transporter 76T at 14.7% (0-58.6%). Additionally, sulfadoxine-pyrimethamine-associated mutations show high frequencies.

Overall, *K13* mutations are rapidly expanding in the region further endangering control efforts with the potential of engendering partner drug resistance.

## Background

Malaria is a life-threatening disease responsible for 249 million cases and 608,000 deaths in 2022, with most cases and deaths attributable to *Plasmodium falciparum* (*Pf*) in Africa.[1,2] The World Health Organization recommended artemisinin combination therapies (ACTs) have provided a highly effective treatment over the past two decades in nearly all African countries.[3,4] Artemisinin partial resistance (ART-R) was first described as prolonged parasite clearance in Cambodia in 2009.[5,6] Mutations in the *Pf Kelch13* (*K13*) gene were determined to be associated with ART-R through *in vitro* selection experiments, followed by validation *in vivo* through field studies and *in vitro* through gene editing.[7–11] The clinically-validated *K13* nonsynonymous mutations are within the propeller region and BTB/POZ domain with limited evidence of delayed clearance associated with variants outside these regions.[12]

K13 561H was the first validated mutation to be described in Africa from samples collected in 2014 from Rwanda.[7] Subsequent assessment in Rwanda has been limited but prevalence appears to be increasing and it has been found in bordering regions of Uganda and Tanzania (ref).[13,14] The emergence of *K13* mutations in East Africa represents a major threat to public health as decreased sensitivity to ART is thought to have expedited partner drug resistance and led to ACT treatment failure in Southeast Asia.[15] Concerningly, other validated mutations, including 675V and 469Y, have been described across the region, as well as in Rwanda albeit at comparatively low prevalences (Table S1).[14,16–19] However, despite the early identification in Rwanda, large-scale surveillance for K13 mutations has not been conducted.

Artemether-lumefantrine (AL) is the most widely deployed ACT in Africa and the predominant treatment in East Africa, followed by artesunate-amodiaquine (ASAQ) (Table S2).[1,20] Rwanda has used AL as the front-line therapeutic for uncomplicated malaria since 2006.[21] AL has remained relatively effective as lumefantrine has never been available as a monotherapy and a true stable lumefantrine resistant phenotype remains unclear.[3,22] Decreased susceptibility to lumefantrine has been correlated to mutations in the *Pf. falciparum* multidrug resistance protein 1 (MDR1) as haplotype N86, 186F and D1246 and wild-type *Pf. falciparum* chloroquine resistance transporter (CRT) K76.[9,23,24] Africa has managed to avoid ACT resistance, with high efficacy intact in regions of high ART-R likely due to continued partner drug efficacy, only reporting delayed clearance in Rwanda, Uganda, Tanzania, and Eritrea, making monitoring efforts vital to inform appropriate responses in these and nearby areas.[12,14,22]

To better characterize the prevalence and dynamics of ART-R and partner drug resistance mutations, geographically broad longitudinal surveillance is needed. Data from Uganda covering 2017 to 2021 is the most robust example of this type of data, but is limited by having one site per province limiting the geographic interpretations (16 sites in a country of 93,065 mi^2^).[13] In addition, the speed by which data is generated is critical for a rapid understanding that can lead to the implementation of control measures. Novel techniques, such as pooling of samples for deep sequencing of target genes, have the potential to increase the resolution and speed of reporting resistance data. For targeted sequencing, previous work has found small and large pools can provide accurate frequency of antimalarial resistance genes.[25,26] To date, only a few studies have used pools to look at either antimalarial resistance or vaccine antigen diversity.[27–30]

Rwanda is a relatively small country (26,338 km^2^) with known ART-R, but lacks dense routine surveillance. We employed a pooling strategy for surveillance across a large geographic area including sites in Rwanda and in neighboring regions of Uganda, Tanzania, and the Democratic Republic of the Congo (DRC) to provide a rapid view of the current state of ART-R across the Great Lake region. To accomplish this, we leveraged a liquid blood drop (LBD) sampling technique, which allowed for rapid pooling and extraction, and high throughput molecular inversion probes (MIPs) targeted to all major known antimalarial resistance polymorphisms.

## Methods

### Sample Collection

Sample collection occurred at 20 health facilities in Rwanda, 6 in Tanzania, 3 in Uganda, and 10 in DRC between May 2022 and March 2023 (Figure 1 and Table S3). Sites in Rwanda were selected in coordination with the National Malaria Control Program to achieve representation of high to medium-prevalence areas as well as those with evidence of increasing transmission based on incidence. Sites in neighboring countries were selected based on detecting potential border traversal and further spread as well as feasibility.

**Figure 1:**
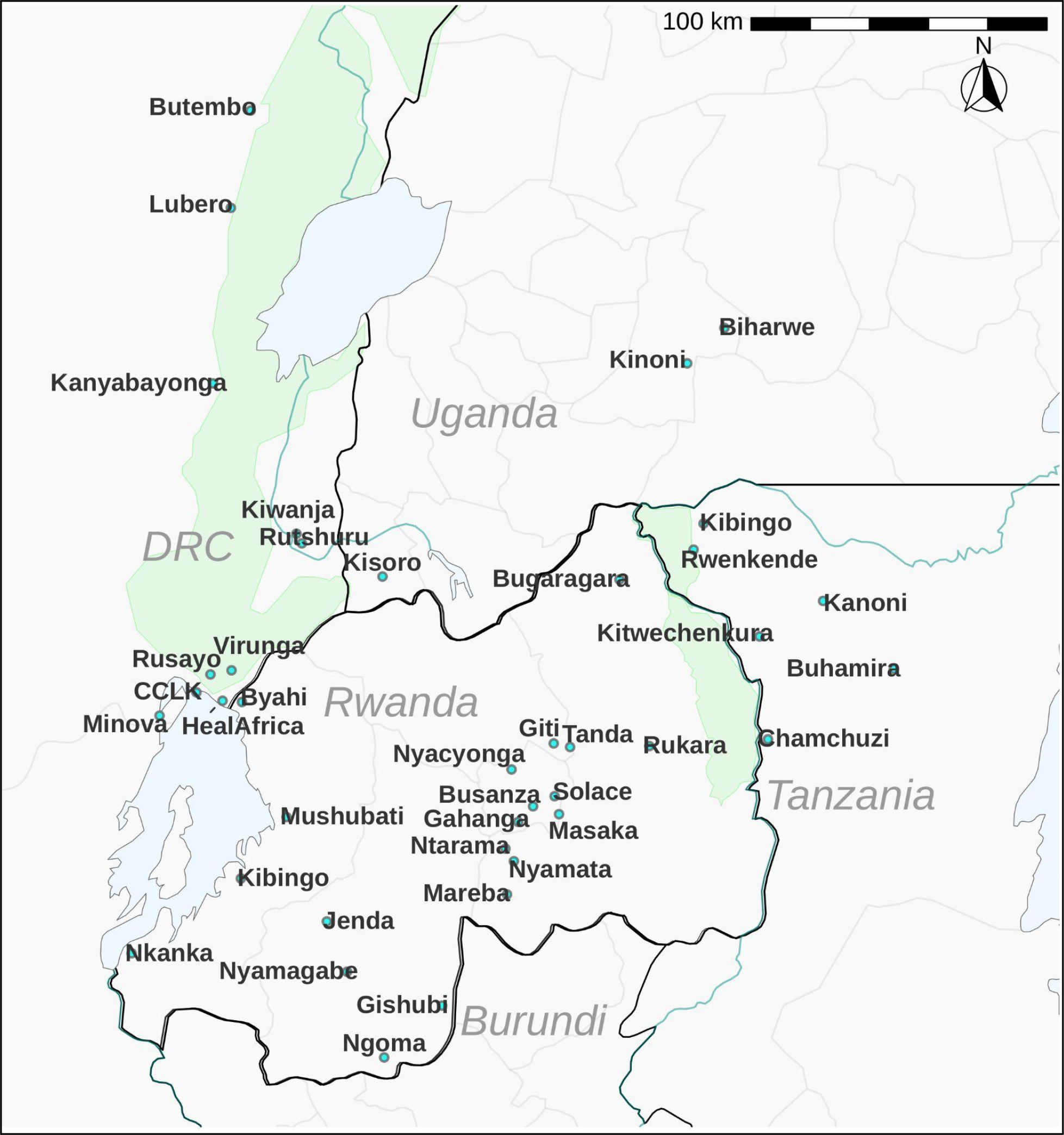
Collection Locations. Map depicting distribution of 39 health centers within Rwanda, Democratic Republic of the Congo, Tanzania and Uganda where health care workers are enrolling and sampling malaria patients after diagnosis.

Malaria-positive patients (n=5,465) were identified by routine microscopy or malaria rapid diagnostic tests (mRDTs). Trained healthcare workers from the facilities consented patients and subsequently collected whole blood, adding 3-4 drops (approximately 100 µl) of capillary blood into a 2mL O-ring microcentrifuge tube pre-filled with 100 µl of 2X DNA/RNA Shield (Zymo Research, Irvine, CA). Collection tubes were vigorously shaken to mix blood with the preservative and stored at room temperature for 2 weeks or less before being transported to institutional −20° C facilities within the country of origin. All samples were then transported to the Center for Genomic Biology (CGB) at INES-Ruhengeri University (Figure 2). Pools were made from individual samples based on collection dates and batch receipt from health care facilities. If a site provided 8 or fewer samples at a given storage date, multiple dates were combined. 6 µl of each sample was pooled into a microfuge tube and vortexed thoroughly before 100 µl was taken for nucleic acid extraction. Pools with insufficient volumes (<=16 samples) were brought to 100 µl using 1x DNA/RNA shield. Details of the pools analyzed are provided in Table S4.

**Figure 2:**
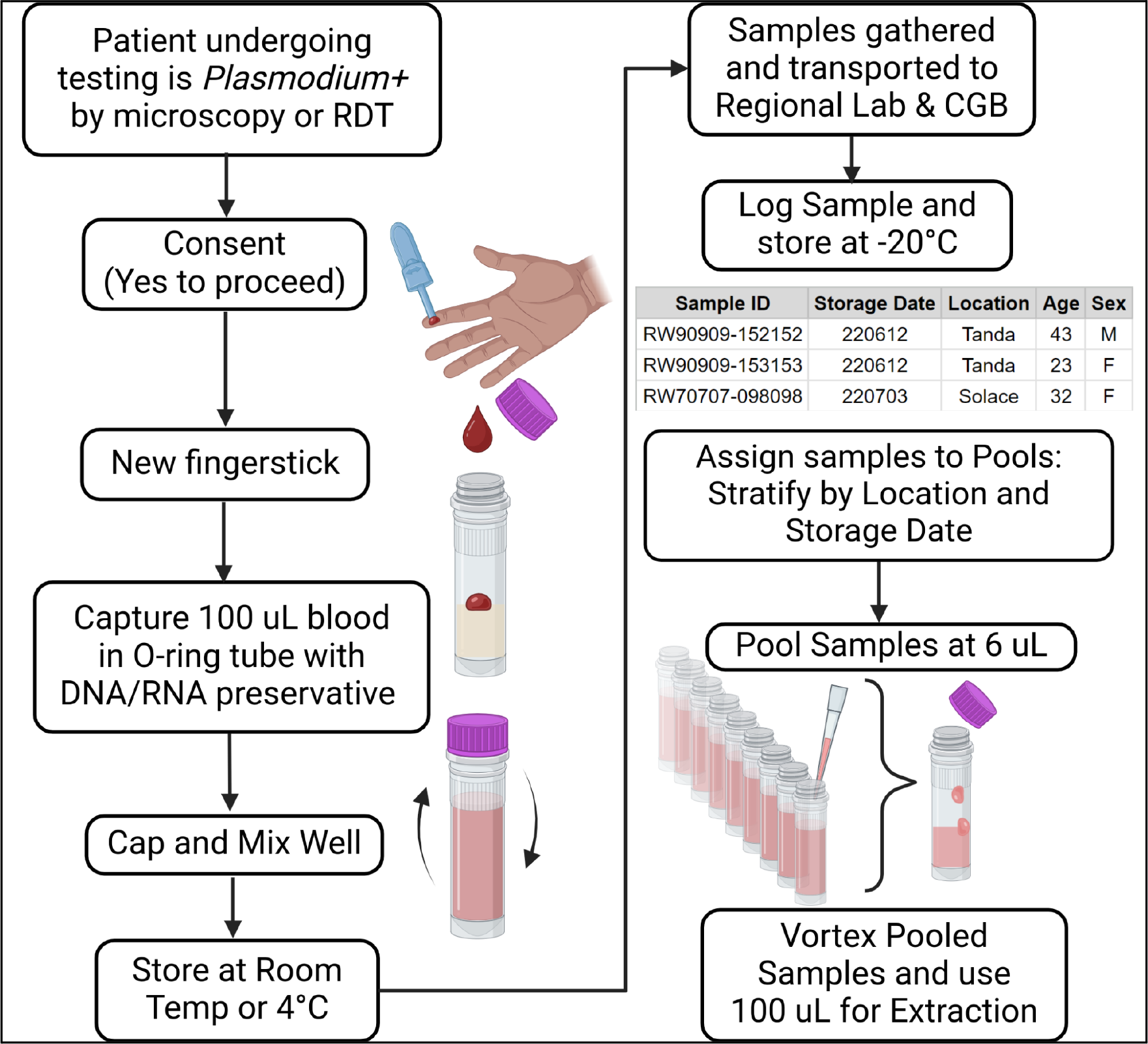
Liquid Blood Drop Collection and Pooling Schema. Workflow describing the patient interaction, sampling, transport and pooling stratification criteria. CGB = Center for Genomic Biology

### Molecular Inversion Probes (MIPs) Genotyping

DNA was extracted and purified using a custom magnetic bead technique (Supplementary Materials). MIP genotyping was performed as detailed in the supplement and by Aydemir et al. using a subset of the MIP panel described as DR2.[31] These 121 probes are associated or suspected to be associated with drug resistance (Table S3, detailed methods supplemental). In brief, MIP capture reactions were performed at Brown University in a 384 well format, followed by exonuclease digestion. A subsequent PCR reaction for each sample, converted the circular MIPs into sequenceable libraries complete with sample barcodes, unique molecular identifiers (UMIs) and Illumina adapters. These individual sample libraries were combined into a single microfuge tube and bead cleaned followed by further purification using agarose gel excision of the properly sized band, which was then sequenced on an Illumina NextSeq 550. If initial sequencing did not achieve sufficient depth were re-pooled, solid phase reversible immobilization (SPRI) bead cleaned, gel extracted, and resequenced. Samples that still did not cumulatively reach the UMI threshold were recaptured and sequenced.

### Analysis

Samples were demultiplexed using previously described software available at https://github.com/bailey-lab/MIPTools.[32] Downstream analysis was performed using R version 4.2.3. UMIs were used to cluster sequences and link amplified single nucleotide polymorphisms (SNPs) occurring from the same probe to properly downsample sequencing data. Frequency data for different polymorphisms were summarized using `ggplot2` and `dplyr` packages in R version 3.6.2 software. Map figures were created in RStudio using spatial data from `rnaturalearth`, `sf` and `ggplot2`. Permutation test of observed candidate mutation presence was determined relative to validated mutation distributions at seed (1979) and 100,000 permutations.

## Results

### Generation of Mutation Frequency Data from Pooling within Sites

Healthcare workers at sites across the 4 countries collected 5,465 samples from May 2022 to March 2023. We generated 199 pools with an average pool size of 27 samples (range: 5-88; median: 25; Table S4). Three rounds of sequencing with library rebalancing yielded sufficient data for 180 pools (90.4%) representing 6.47e7 total reads with 102/104, 52/55, 24/35, and 2/5 successful pools from Rwanda, Tanzania, DRC, and Uganda, respectively. This represented 20/20, 6/6, 8/10, and 1/3 site(s) from Rwanda, Tanzania, DRC and Uganda, respectively. Unsuccessful pools correlated to low DNA yields by Qubit and correlated with earlier collection in the study when a fraction of samples contained clots due to overfilling and inadequate mixing upon collection, which was resolved with further healthcare worker training. We calculated the site resistance allele frequency as an average of all pools within a site, weighted by the number of samples per pool along with the average of the sites (equally weighted) across a country (Table 1). Sites lacking sequence data were excluded from analysis.

**Table 1:** Average site frequencies of WHO Validated and Candidate *K13* Mutations.

|  | E. DRC | Rwanda | NW. Tanzania | SW. Uganda |
| --- | --- | --- | --- | --- |
| Sites | 8 | 20 | 6 | 1 |
| Mutation | Avg Freq (Range) | Avg Freq (Range) | Avg Freq (Range) | Avg Freq (Range) |
| **Validated** |  |  |  |  |
| 469Y | 0% (0-0%) | 0% (0-0%) | 0.1% (0-0.7%) | 0% (0-0%) |
| 561H | 0% (0-0%) | 19% (0-54.5%) | 12.1% (0-35.2%) | 1.6% (1.6-1.6%) |
| 574L | 0% (0-0%) | 0.7% (0-8.8%) | 0.3% (0-1.8%) | 0% (0-0%) |
| 675V | 0.8% (0-4.9%) | 5% (0-35.5%) | 0.1% (0-0.5%) | 0% (0-0%) |
| **Candidate** |  |  |  |  |
| 441L | 0% (0-0%) | 0.8% (0-7.7%) | 3.8% (0-17.2%) | 1.8% (1.8-1.8%) |
| 449A | 0% (0-0%) | 1% (0-12.8%) | 0% (0-0%) | 0% (0-0%) |
| 469F | 0% (0-0%) | 2.7% (0-42.6%) | 0.1% (0-0.5%) | 4.5% (4.5-4.5%) |
| 568G | 0% (0-0.2%) | 0% (0-0.6%) | 0% (0-0%) | 0% (0-0%) |

\*E. DRC = Eastern Democratic Republic of the Congo, NW. Tanzania = Northwestern Tanzania, SW. Uganda = Southwestern Uganda

### Multiple validated K13 mutations are found in the region with 561H showing greatest frequency and geographic distribution

K13 561H reached an overall average site frequency in Rwanda of 19.0% and is found in 90% (18/20) of Rwandan sites surveyed, with the highest frequency in Kibingo at 54.5% (Table 1 and Figure 3). Higher 561H frequencies clustered near Kigali City and it was the dominating K13 mutant in 15 of 18 sites (Figure 3). 561H was also detectable in 5 sites from Tanzania and the sole sequenced site in Uganda. The average site frequency in Tanzania was 12.1%; the highest single facility was Rwenkende in the North (35.2%, Figure 3). There was no 561H detected in the DRC pools, representing 566 samples.

**Figure 3:**
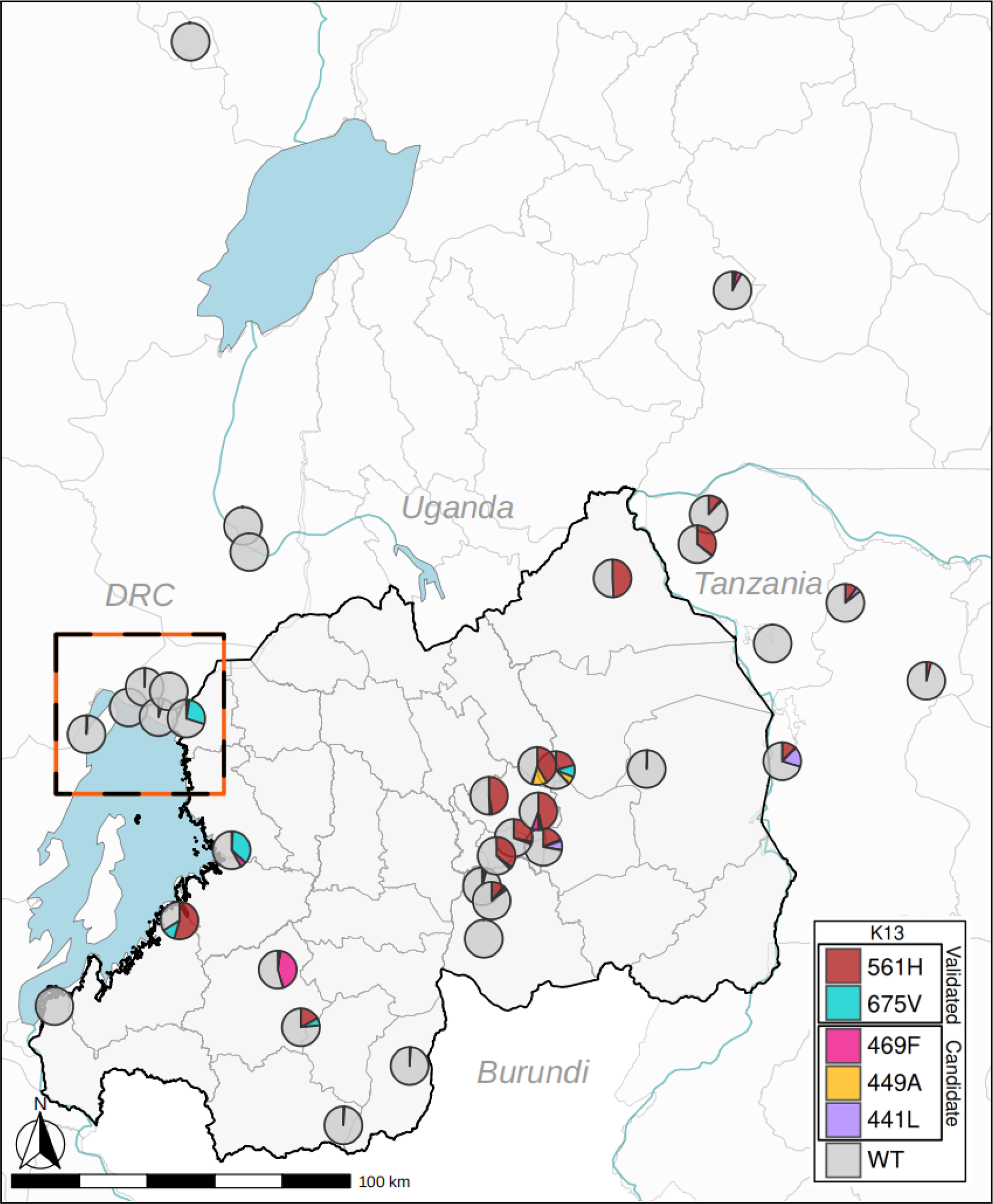
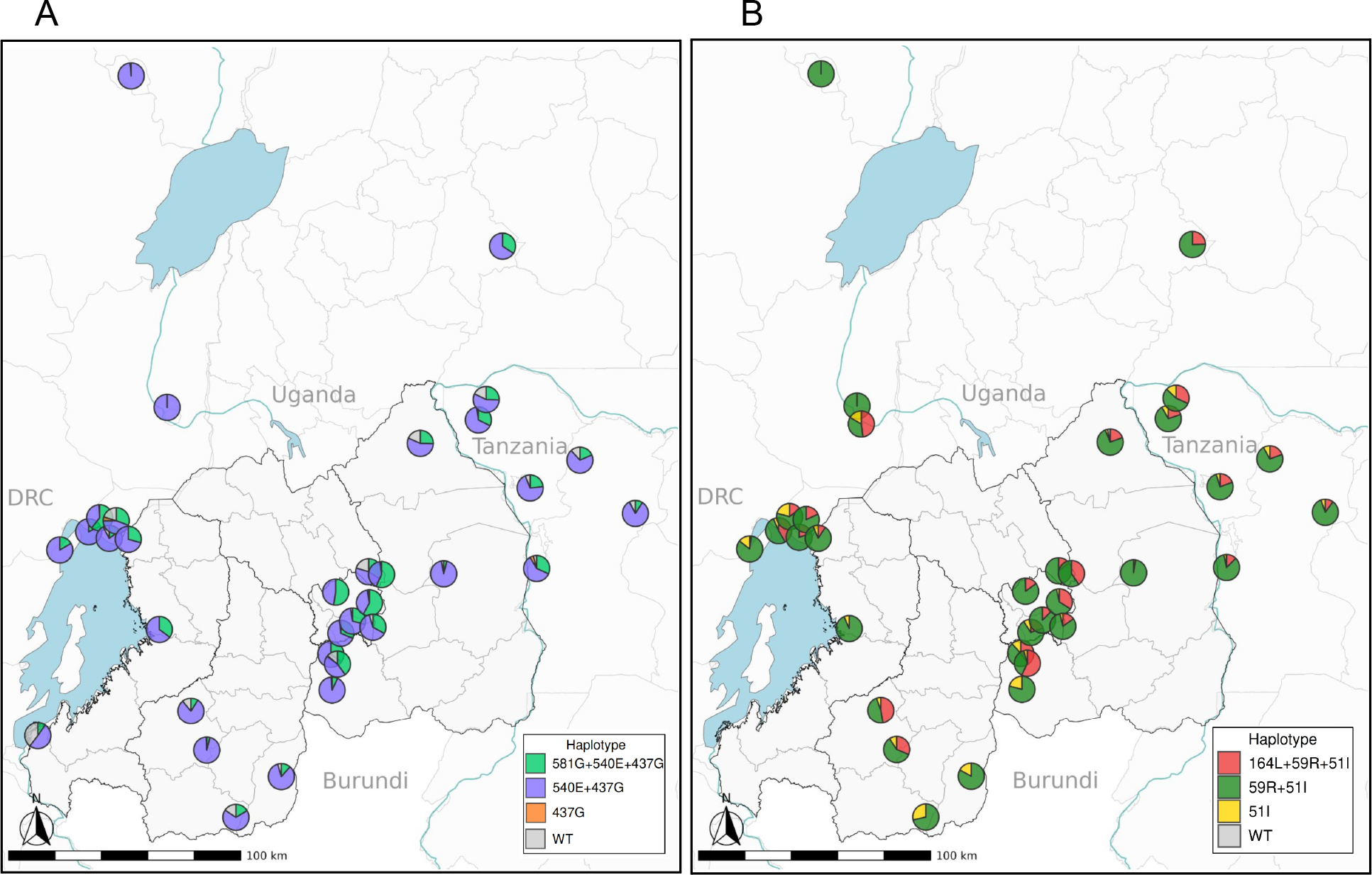
Spatial Distribution of the most frequent WHO mutations detected in Rwanda and neighboring countries. Geographic distribution for the highest frequency WHO validated and candidate mutations in Rwanda and neighboring countries. Average site frequencies and location of clinics are represented by pie graphs with colors representing individual validated and candidate mutations. Black and orange dashed rectangle highlights neighboring regions of DRC and Rwanda with disparate K13 mutation frequencies. A) Distribution *DHPS* mutation haplotypes (imputed from frequency data) B) Distribution *DHFR* mutation haplotypes (imputed from frequency data) We impute haplotypes based on the order of appearance of these mutations. In the case of *DHPS*, 437G appeared first and is at high frequency so any remaining wildtype A437 is charted in grey. For the mutant portion of the pie chart, the frequencies were adjusted to present the ratio of mutant 540E and 581G, if these two frequencies did not total the frequency of 437G, then the remaining portion is labeled at 437G.

Despite occasional low-level detection in previous reports from Rwanda, 675V appears geographically widespread, albeit at relatively lower frequency (overall average frequency of 5.0%) and detection in 13 of 20 sites. The site frequency was higher along Lake Kivu, being greatest at Mushubati (35.5%) (Figure 3). 574L was the only other validated WHO mutation, found at 3 sites in Rwanda, with an average site frequency of 0.7% and greatest frequency at Nyamagabe (8.8%). In neighboring countries, 675V was detected in DRC at 3 sites with an average site frequency of 0.8% as well as at 2 sites in Tanzania with an average site frequency of 0.1%. As for other validated mutations, 469Y was detected at one site in Tanzania.

Within the 20 Rwandan sites, three candidate mutations of concern were detected, with a country site average frequency for 441L of 0.8% detected in 7 sites, for 449A of 1.0% detected in 3 sites, and for 469F of 2.7% detected in 6 sites (Table 1, Figure 3). The only candidate mutation detected in DRC was 568G, from 1 site in a single pool. Tanzania and Uganda sites showed 441L and 469F mutations at frequencies (<5%). MIP data and all detected *K13* polymorphisms are provided in Table S4 and Table S6.

Apart from WHO-recognized mutations, there were 6 other mutations detected in the propeller region, 3 mutations detected within the POZ domain, and 12 mutations detected in the upstream region (Table S6). Of the propeller mutations, 667R reached the highest average site frequency at 1.7% (Table S6). 555A, a mutation historically found within Rwanda,[7,8] was most widely distributed, with detection in 8 sites with the highest site being 8.0% (Table S7).

### Validated and candidate mutations show spatial association in occurrence

Validated and candidate mutations were detected together in 16 sites with no sites exclusively harboring candidate mutations (P=0.0002, permutation test; Methods) (Figure 4). Within the validated mutations, 11 sites had both 675V and 561H. 469Y and 574L did not occur independently in any sites (Table S8). Among candidate mutations 469F and 441L were the most common co-occurring mutations (Table S7).

**Figure 4:**
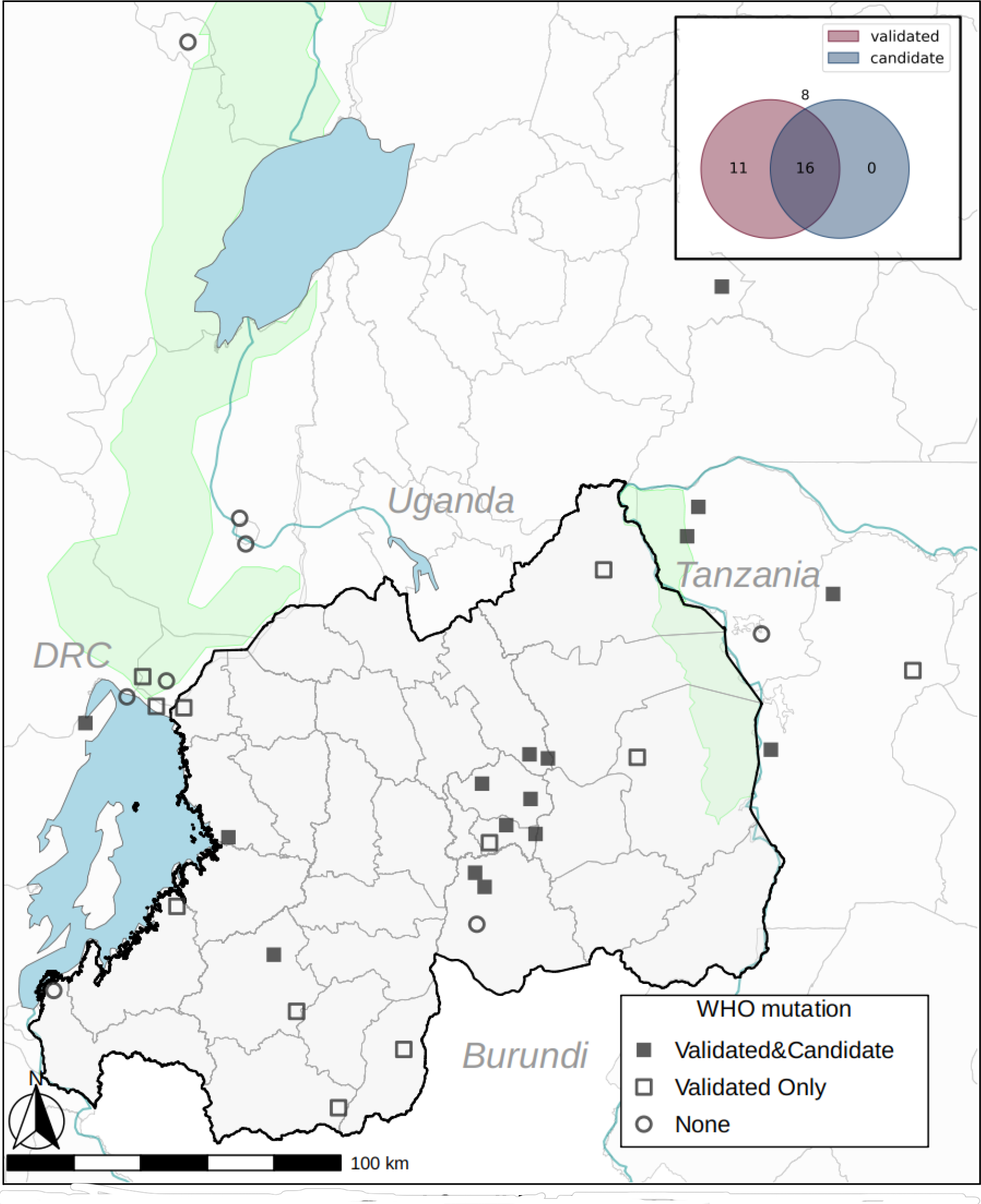
Patterns of Validated and Candidate Mutation Co-occurrence. A) Site counts of validated and candidate mutation Co-occurrence. B) Co-occurrence of validated mutations C) Co-occurrence of candidate mutations among 35 sequenced sites

### Disparate mutation frequencies between neighboring sites in DRC and Rwanda

We detected large frequency differences for 561H and 675V at the border between Rwanda and the DRC border. The province weighted average frequency for 561H was 14.5% and 0% for the Western Province, Rwanda and Goma District, DRC, respectively (Figure 3b). Similarly, the province weighted average frequency for 675V was 18.6% and 1.7% for the Western Province, Rwanda and Goma district, DRC, respectively. At Byahi, Rwanda, we detected a 561H frequency of 2.7% and a 675V frequency of 27.2% in a pool of samples from Oct-Dec 2022. Meanwhile, at a site 6.5km away in Goma, DRC, pools from a month prior (Sept 2022), we detected no 561H mutations and only a frequency of 4.9% for 675V. This discrepancy in K13 mutation frequency between sites in DRC and Rwanda is occurring despite a high degree of human migration, with nearly 80,000 people normally crossing the DRC-Rwanda border per day in this region.[33] We did detect low 675V frequencies in two other DRC health facilities - Minova (1.0%) and Rusayo (0.3%) - more distant from the border earlier in the year (Aug 2022 and July 2022, respectively).

### Resistance mutations associated with other antimalarials are widely distributed through the Great Lakes region

Table 2 summarizes average site frequency estimates for major antimalarial resistance polymorphisms. In terms of ACT partner drug resistance, mutations in MDR1 that affect tolerance to lumefantrine were widely distributed in the region, as well as the wild type alleles that are part of the NFD haplotype (Table 2). Average overall frequency of 184F was 47% with the highest site presenting nearly 95% frequency (Figure 5).

**Table 2:** Partner Drug Resistant Mutations.

|  | **E. DRC** | **Rwanda** | **NW. Tanzania** | **SW. Uganda** |
| --- | --- | --- | --- | --- |
| Mutation | Avg Freq (Range) | Avg Freq (Range) | Avg Freq (Range) | Avg Freq (Range) |
| **CRT** |  |  |  |  |
| M74I | 58.8% (16.5-95.8%) | 14.7% (0-58.6%) | 0.6% (0-3.9%) | 0% (0-0%) |
| K76T | 58.8% (16.5-95.8%) | 14.7% (0-58.6%) | 0.6% (0-3.9%) | 0% (0-0%) |
| Q271E | 48.5% (8.4-100%) | 19.7% (0-71.7%) | 3.3% (0-11.5%) | 0% (0-0%) |
| R371I | 51.9% (8.3-94.6%) | 16.4% (0-69.3%) | 3.4% (0-8.9%) | 0% (0-0%) |
| **DHFR** |  |  |  |  |
| N51I | 100% (100-100%) | 99.8% (96.7-100%) | 99.9% (99.7-100%) | 100% (100-100%) |
| C59R | 92.7% (79-100%) | 92.6% (71.5-100%) | 92.9% (87.1-96.6%) | 100% (100-100%) |
| S108N | 100% (100-100%) | 99.8% (95.2-100%) | 100% (100-100%) | 100% (100-100%) |
| I164L | 19.1% (0-48%) | 18.1% (0-57.1%) | 18.9% (10.1-31.7%) | 32.5% (32.5-32.5%) |
| **DHPS** |  |  |  |  |
| A437G | 95.8% (79.7-100%) | 93.1% (60.5-100%) | 91.7% (82.1-97.8%) | 100% (100-100%) |
| K540E | 95.3% (74.8-100%) | 95.3% (81.8-100%) | 91.3% (82.6-98.4%) | 100% (100-100%) |
| A581G | 21.3% (0-60.7%) | 33% (0-64.4%) | 30.3% (9.1-48.6%) | 52.2% (52.2-52.2%) |
| **MDR1** |  |  |  |  |
| N86Y | 17.3% (0-95.8%) | 2% (0-36.7%) | 0% (0-0%) | 0% (0-0%) |
| Y184F | 30.6% (0-56.9%) | 47% (0-94.3%) | 45.1% (29.1-49.7%) | 87.1% (87.1-87.1%) |
| F938Y | 9.4% (0-37.4%) | 1.3% (0-9.1%) | 0% (0-0%) | 0% (0-0%) |
| N1042D | 0.9% (0-3%) | 0% (0-0%) | 0% (0-0%) | 0% (0-0%) |
| D1246Y | 28.7% (0.6-97.8%) | 6.1% (0-37.9%) | 0.5% (0-1.5%) | 3.1% (3.1-3.1%) |

\*E. DRC = Eastern Democratic Republic of the Congo, NW. Tanzania = Northwestern Tanzania, SW. Uganda = Southwestern Uganda

**Figure 5:**
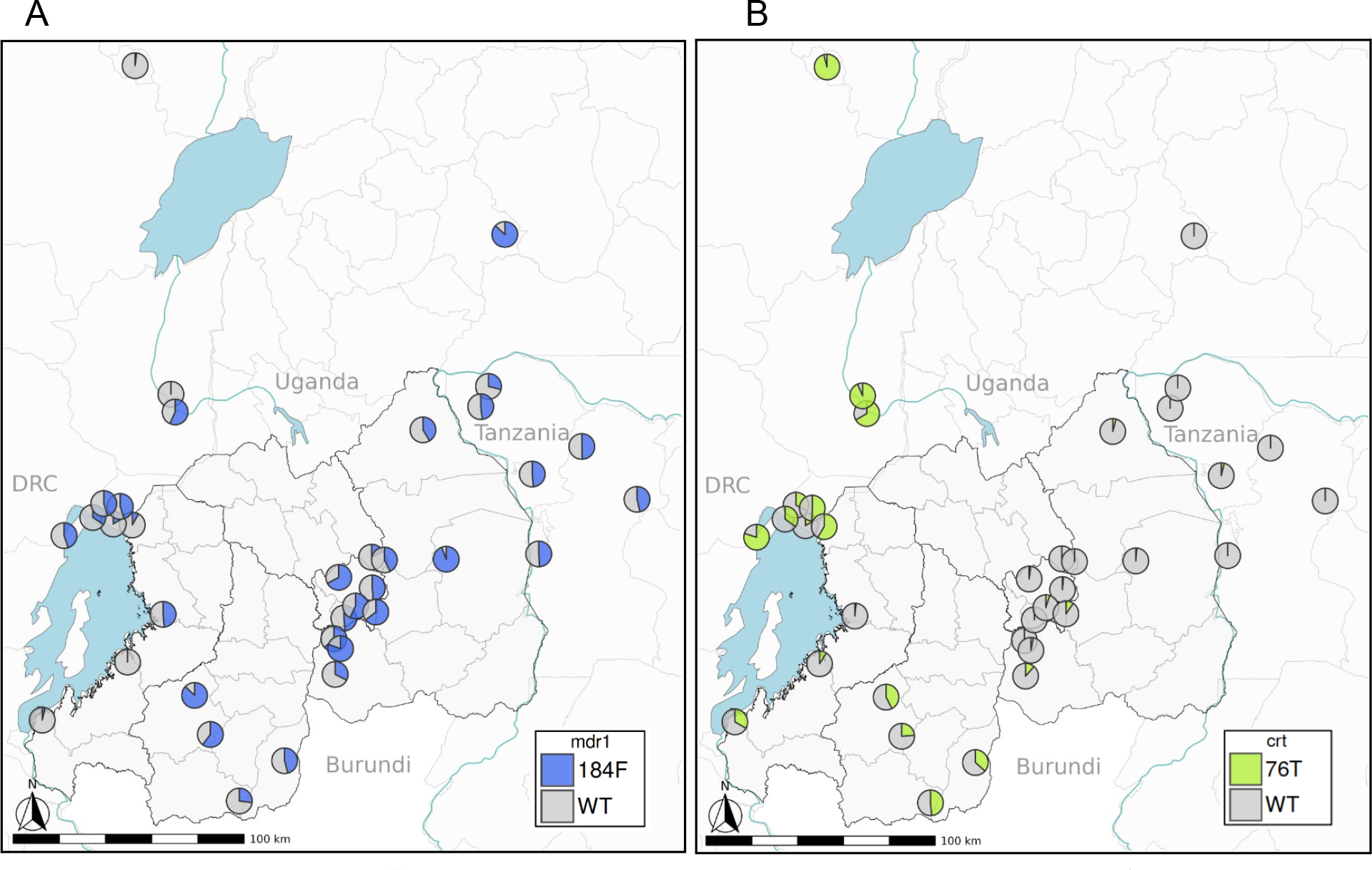
Partner Drug Resistant Mutations. A) Depicts geographic distribution of frequencies found of MDR1 184F according to collection facility pools were sourced from. Frequencies are available in supplemental table 4. B) Depicts geographic distribution of frequencies for CRT 76T.

Mutations in CRT associated with amodiaquine resistance were highest in DRC where ASAQ is used (Table 2), but also detected appreciable frequencies in Rwanda, but with a clustering near the border with Burundi (Figure 5).

In terms of other antimalarials used in the region (Table S2), sulfadoxine-pyrimethamine (SP) continues to be used in the countries surrounding Rwanda. Mutations associated with high level SP resistance, such as dihydrofolate reductase (DHFR) 164L and dihydropteroate synthase (DHPS) 581G were commonly found (Table 2) and widely distributed including in Rwanda where IPTp is not routinely used (Figure S1).

## Discussion

After early reports of *K13* mutations in Rwanda, there has been little investigation into the prevalence and geographic distribution of *K13* mutations. We used pooled sequencing of liquid-preserved blood drops to more rapidly and cost-effectively surveil numerous samples across a larger number of sites compared to traditional dried blood spots. Our findings, derived from a more representative national sample of medium and high-transmission areas, demonstrate widespread K13 561H across 18 of 20 sites in Rwanda, at an overall weighted frequency of 19%, but ranging upwards of 54.5%.

Notably, we also found evidence of 561H in neighboring countries including Tanzania and Uganda, albeit at lower relative frequencies. Together, these results highlight not only the value of rigorous, trans-national surveillance programs, but also robust proof-of-concept for LBD collections and pooled targeted sequencing to provide timely and accurate molecular information to guide control efforts.

Interesting patterns of allele frequencies in ART-R and partner drug resistance polymorphisms were noted across each of the bordering countries, highlighting the complex nature of resistance emergence and spread. Along the DRC-Rwanda border, we see a stark differentiation of ART-R allele frequencies across the border. The most likely explanation for this is the differing drug policies between the countries, with AL use in Rwanda and more ASAQ use in DRC. This is supported by the contrasting allele frequencies in mutations that are oppositely selected by the partner drugs (MDR1 and CRT). We note more mutant MDR1 86Y in DRC and more 184F in Rwanda, Uganda and Tanzania, consistent with ASAQ and AL usage policy in the region. We however see a dissimilar pattern at the border with Burundi, where CRT polymorphisms are higher in bordering Rwanda areas than in other regions. Like DRC, Burundi used ASAQ, but on the DRC border we do not see increased allele frequencies in *Pfcrt* mutations as widely distributed on the Rwanda side. Put together, these data suggest that the relationship dictating cross border movement of antimalarial resistance is likely highly complex. All of these borders have high rates of human mobility and geographic proximity, but differ in the antimalarial resistance profiles that can not fully be explained by antimalarial policies alone. This suggests that other factors, potentially such as transmission intensity, may play a role in these relationships. Given resistance to ACTs requires mutations in multiple genes to affect both drugs, differences in transmission may provide a natural barrier to limit spread.[34] More detailed studies of transmission leveraging genomics and vector biology could help elucidate these underlying mechanisms.

We provide all evidence of additional kelch polymorphism and the distribution of DHPS and DHFR mutations using a timeline imputation to report haplotypes in Figure S1. We saw DHPS 581G mutations in 19 sites within Rwanda reaching an overall weighted average of 33.0%. DHFR 164L was in 17 sites, reaching a country average of 18.1% (Table S9). Similar to reports of mutant DHFR/DHPS in Tanzania[18], all sites in this study saw at least two mutant SNPs of DHPS and DHFR; these data raise concern for general use of SP in perennial malaria chemoprevention.

On the Tanzania border, we also see 561H reaching a high frequency. Unlike the DRC, Tanzania uses the same first line antimalarial, AL, as Rwanda. This may have facilitated the movement of 561H mutations between Rwanda and Tanzania that was not possible on the DRC side. In addition, there may have been an independent origin of 561H within the bordering regions of Tanzania.[18] 561H is becoming ubiquitous across the Rwanda, the 675V mutation was seen at an average frequency of 5% across the country, with the highest site frequencies occurring along Lake Kivu. The other validated mutation detected, 574L, was found at a single site at 8.8%. Additionally, candidate mutations 441L, 449A, and 469F, were detected in Rwanda dispersed in 11 sites at low frequencies. Another striking finding is the geographic relationship between validated and candidate ART-R polymorphisms. We failed to detect any candidate polymorphisms in sites where there were no validated ART-R polymorphisms. This suggests candidate polymorphisms are likely advantageous in similar places where established resistance is appearing, given that validated mutations arise from drug pressure, these candidate mutations are also likely taking advantage of similar selection pressure.

Our methods of large scale sampling and pooling created a major advantage in terms of speed, but were inherently subject to limitations. Early sample collections within the first weeks were inadequately mixed, resulting in clotted blood which yielded poor DNA extraction and subsequent sequencing failure. This laxity was partly due to the wide geographic area in our study and gaps in monitoring and training were addressed in later months. Pooling samples allowed us to sequence thousands of samples in 3 months and save reagents, but this method obfuscates any complexity of infection, haplotype or identity-by-descent analysis that could provide information about mutation spread. It appears that partial resistance in Rwanda has risen to a level where pooling samples is only effective as an initial assessment, individual level results can incorporate more refined geographic data, and provide detailed study of each district.

This represents one of the most geographically dense sampling surveys of ART-R resistance mutations and provides a detailed picture of one of the epicenters of emergence in Africa. The broad geographic range affected by the presence of these mutations is of significant concern for malaria control in the country and potentially for neighboring countries. Recent modeling research has suggested that ASAQ should be substituted for AL to slow treatment failure.[35] We found that sites with higher *K13* mutant parasite frequencies corresponded to low ASAQ resistance markers. More research to understand the fitness of circulating mutant strains under ASAQ pressure is needed. Our study data demonstrating that 561H and other *K13* mutations are now widely distributed across Rwanda and into neighboring Tanzania and Uganda provides a necessary update to improve modeling and understand current conditions.

## Conclusion

We highlight that since initial reports from samples collected in 2014, ART-R polymorphisms now are at high frequency in 2022 samples with a broad geographic distribution across Rwanda. We document that K13 561H is a national problem and that other ART-R mutations are present and at significant frequencies in certain regions. We see a complex relationship of allele frequencies across differing borders that is partially explained by drug policy, but likely impacted by other unmeasured factors. Lastly, the relationship between validated and candidate ART-R polymorphisms by geography suggests a complicated relationship concerning initial emergence of resistance. Overall, these data are highly concerning for the longevity of ACTs in Africa, and potentially set the stage for the emergence of partner drug resistance that can lead to the collapse of ACT efficacy. In addition, it is clear that additional studies are required to help us better understand the emergence of resistance and all the factors that may impact the spread across national borders.

## List of Abbreviations

(Pf): Plasmodium falciparum
(ACTs): artemisinin combination therapies
(ART): artemisinin
(ART-R): ART partial resistance
(K13): Plasmodium falciparum Kelch13
(AL): Artemether-lumefantrine
(ASAQ): artesunate-amodiaquine
(MDR1): P. falciparum multidrug resistance protein 1
(CRT): P. falciparum chloroquine resistance transporter
(DRC): Democratic Republic of the Congo
(LBD): liquid blood drop
(MIPs): molecular inversion probes
(mRDT): malaria rapid diagnostic test
(CGB): Center for Genomic Biology
(SPRI): solid phase reversible immobilization
(UMI): Unique Molecular Identifiers
(SNPs): single nucleotide polymorphisms
(SP): sulfadoxine-pyrimethamine
(DHFR): dihydrofolate reductase
(DHPS): dihydropteroate synthase

## Declarations

### Ethics approval and consent to participate

Ethical approval for this study was obtained from the Rwanda National Ethics Committee (reference 12/RNEC/2022), the Medical Coordinating Committee of the National Institute for Medical Research Tanzania, the MUST Research Ethics Committee, Ethics Committee of Ecole de Santé Publique de Kinshasa and the Institutional Review Board at Brown University (RI, USA). University of North Carolina approved this work under a reliance agreement with Brown University. Written informed consent was obtained from participants or parents/guardians of eligible children in the local language which is Kinyarwanda in Rwanda, Kiswahili in Tanzania, French in DRC, and English in Uganda.

## Consent for publication

“Not applicable”

## Availability of data and materials

Illumina data is available from the SRA (Project # pending). All sequenced MIP counts are included within the supplementary materials. MIPWrangler and MIPTools software is available on GitHub https://github.com/bailey-lab. R scripts used to generate analysis and figures are available at https://github.com/nnwyoung/ArtR_Pooled_MIPs.

## Conflict of Interest/Competing Interests

The authors have no conflicts of interest to report. The funder had no role in the implementation and interpretation of the project.

## Funding

This project was funded by the National Institute of Allergy and Infectious Diseases (R01AI156267 to JAB, JJJ, DSI, SH, and JBM; K24AI134990 to JJJ).

## Authors’ contributions

NWY contributed to the design of the work, data collection, laboratory procedures, analysis, interpretation of results, drafting, and revision of the manuscript. GP, TM, FM, FM, RB, RMB, CIM, GKK, EM, DSI, and SH contributed to the design of the work and data and specimen collection. RC, AL, VI, and NJB contributed to laboratory procedures and data collection. DG, JBM, and CK contributed to the design of the work and revision of the manuscript. JJJ and JAB contributed to the design of the work, interpretation of results, and revision of the manuscript.

## Acknowledgements

The authors extend thanks to all participants involved for contributing to this study. We thank the staff from all 39 health facilities for their willingness to help in obtaining liquid blood drops and demographic information. The support from colleagues at NIMR - Tanzania (Misago Seth, Rashid Madebe, Tilaus Gustav, Ildephonce Mathias,Gerion Gaudin, Christopher Masaka, Neema Manumbu, and Millen Meena) is very much appreciated. This work would not be possible without various bus drivers and other transportation experts. We thank them for their help in delivering materials to neighboring countries.

## Authors’ information

This section is optional.

## Supplementary Materials

## Supplementary Tables

**Table S1.** Previous Reports of Validated Artemisinin Partial resistance Mutations in Rwanda.

**Table S2:** Antimalarial Policies in 2022 for *Plasmodium falciparum* by Country (Adapted from WMR 2023)[1].

**Table S3:** Sample_Collection.

**Table S4a-d:** Rwanda_UMI_counts, DRC_UMI_counts, UG_UMI_counts, TZ_UMI_counts.

**Table S5:** DRexMini_MIP_panel (in xlsx) Table S6: Other *K13* polymorphisms (in xlsx)

**Table S7:** Average Frequencies by Health Facility.

**Table S8:** Health Facility Occurrence Counts by Country Table S9: Average Frequencies by Country.

## Supplementary Figures

**Figure S1:** Partner Drug Resistance Haplotypes

## Supplementary Methods

### DNA Extraction

Pools were spun down and combined with 100 µl Lysis Buffer (80 mM DTT, 2M Guanidine thiocyanate, 25 mM Sodium Citrate Dihydrate, 0.1 M Tris HCl, Proteinase K, and DiH_2_O). The sample was incubated at 65° C for 10 minutes and allowed to cool before the addition of 3 µl of RNase A (Invitrogen, Carlsbad CA, USA), 400 µl of absolute ethanol, and 15 µl of Spherotech SPHERO Silica Super Paramagnetic Beads (Lake Forest, IL) with mixing between each addition. Samples were mixed by rotation for 10 minutes and then placed on a magnet. The supernatant was removed and the paramagnetic beads were washed twice with 80% ethanol. After the last wash, the sample was removed from the magnet, allowed to dry briefly, then eluted with 60 µl of 1X TE Low EDTA (Thermo Fisher Scientific, Waltham MA, USA) by heating to 65° C for 10 minutes and then returned to the magnet. Once beads were pelleted to the magnet, the elution was removed and a random subset was quantified with Qubit Fluorometric Quantification (Thermo Fisher Scientific, Waltham, MA, USA) as a quality check.

Extracted DNA was subsequently stored at −20° C to await MIP genotyping.

### Library Preparation

After the PCR addition of barcodes and adapters, 5 uL of all pools were combined into a single tube and cleaned prior to sequencing to remove lower-weight non-specific PCR products. To prepare libraries for sequencing, a 1.2x SPRI bead cleanup was performed on the final library and eluted in 50 µl TE low EDTA pH=8 (Thermo Fisher Scientific, Waltham MA, USA). The amplicons were resolved on a 1.0% agarose gel. DNA fragments of the correct size were excised and purified using the NEB DNA Gel

Extraction Kit T1020S (Ipswich, MA, USA). Library quality was assessed using a fragment analyzer (Agilent, Santa Clara, CA), the library was pooled with other libraries, denatured with 0.2N Sodium Hydroxide, spiked with 2.5% Phi-X and sequenced on an Illumina Nextseq 550 (Illumina, San Diego, California) using a 300-cycle mid-output kit based on the manufacturer’s instructions.

### Re-pooling

Samples that did not achieve >=1,000 Unique Molecular Identifiers (UMI) for the main target mutation (561H) on the first pass of sequencing were re-pooled, SPRI cleaned, gel extracted, and resequenced. Samples that still did not cumulatively reach the UMI threshold were recaptured and sequenced.

## Notes

### Competing Interest Statement

The authors have declared no competing interest.

